# Large-scale identification of social and behavioral determinants of health from clinical notes: Comparison of Latent Semantic Indexing and Generative Pretrained Transformer (GPT) models

**DOI:** 10.1101/2024.04.22.24306142

**Authors:** Sujoy Roy, Shane Morrell, Lili Zhao, Ramin Homayouni

**Author notes:** **Correspondence:** Ramin Homayouni, PhD, Professor, Foundational Medical Studies; and Director, Population Health Informatics Oakland University William Beaumont School of Medicine, 586 Pioneer Dr, 460 O’Dowd Hall, Rochester, Michigan, 48309-4482, United States of America.

## Abstract

**Background:** Social and behavioral determinants of health (SBDH) are associated with a variety of health and utilization outcomes, yet these factors are not routinely documented in the structured fields of electronic health records (EHR). The objective of this study was to evaluate different machine learning approaches for detection of SBDH from the unstructured clinical notes in the EHR.

**Methods:** Latent Semantic Indexing (LSI) was applied to 2,083,180 clinical notes corresponding to 46,146 patients in the MIMIC-III dataset. Using LSI, patients were ranked based on conceptual relevance to a set of keywords (lexicons) pertaining to 15 different SBDH categories. For Generative Pretrained Transformer (GPT) models, API requests were made with a Python script to connect to the OpenAI services in Azure, using gpt-3.5-turbo-1106 and gpt-4-1106-preview models. Prediction of SBDH categories were performed using logistic regression model that included age, gender race and SBDH ICD-9 codes with a natural cubic spline of 2 degrees of freedom for age.

**Results:** LSI retrieved patients according to 15 SBDH domains, with an overall average PPV *≥* 83%. Using manually curated gold standard (GS) sets for nine SBDH categories, the macro-F1 score of LSI (0.74) was better than ICD-9 (0.71) and GPT-3.5 (0.54), but lower than GPT-4 (0.80). Due to document size limitations, only a subset of the GS cases could be processed by GPT-3.5 (55.8%) and GPT-4 (94.2%), compared to LSI (100%). Using common GS subsets for nine different SBDH categories, the macro-F1 of ICD-9 combined with either LSI (mean 0.88, 95% CI 0.82-0.93), GPT-3.5 (0.86, 0.82-0.91) or GPT-4 (0.88, 0.83-0.94) was not significantly different. After including age, gender, race and ICD-9 in a logistic regression model, the AUC for prediction of six out of the nine SBDH categories was higher for LSI compared to GPT-4.0.

**Conclusions:** These results demonstrate that the LSI approach performs comparable to more recent large language models, such as GPT-3.5 and GPT-4.0, when using the same set of documents. Importantly, LSI is robust, deterministic, and does not have document-size limitations or cost implications, which make it more amenable to real-world applications in health systems.

## Background

There is growing evidence that Social and Behavioral Determinants of Health (SBDH) are associated with a wide variety of health outcomes and that including SBDH data can improve prediction of health risks. ^1,2^ While many studies focus on using neighborhood level SBDH indicators, evidence suggests that using individual-level SBDH significantly improves prediction of outcomes such as medication adherence, risk of hospitalization, HIV risk, suicide attempts, or the need for social work. ^1^ In contrast, most studies that used external neighborhood-level data showed minimal contribution to individual risk prediction. ^1^ Currently, documentation of individual-level SBDH is sparse and incomplete in the structured fields within the EHR, ^3^ but there are increasing efforts to implement screening tools in clinical workflow to document patient-level SBDH factors. ^4^ However, screening tools add a significant burden on the healthcare staff at a time when provider burnout is a major concern. ^5^

SBDH topics may arise during informal communications between the patient and healthcare provider, which are often documented in the clinical notes rather than the structured fields in the EHR. ^5^ As an alternative strategy to screening questionnaires and diagnosis codes, several groups have evaluated SBDH documented in the clinical notes in the EHR. Navathe et al. reported that the highest rates of social characteristics were found in physician notes and that the frequency of six out of the seven social characteristics increased when comparing data from physician notes with billing codes. ^6^ Similarly, in a larger study, Hatef et al. reported that the prevalence of SBDH in notes was vastly higher compared to billing codes for social isolation (2.59% vs 0.58%), housing issues (2.99% vs 0.19%), and financial strain (0.99% vs 0.06%). ^7^

Recent work has focused on developing natural language processing (NLP) and machine learning approaches to extract or infer SBDH from clinical narratives. ^8,9^ NLP approaches are rule-based and identify SBDH lexicons (keywords and/or phrases) using keyword matching or regular expressions. Identification of SBDH lexicons and NLP rules require considerable manual refinement. ^10,11^ More recently, supervised machine learning approaches have been explored for identification of SBDH from notes, by combining a variety of embedding methods, such as bag-of-words, n-grams, wod2vec or Bi-directional Encoder Representation from Transformers (BERT), with supervised classification methods such as support vector machines, random forests, logistic regression, convolutional neural network and feed-forward neural network methods. ^8^ More recent methods that combine transformer-based embeddings learned from large volumes of documents (Large Language Models, LLM) and deep learning classifiers have demonstrated superior performance in extracting SBDH from clinical notes. ^12–15^ However, these models require training large amount of external data sources and fine-tuning using positive and negative gold standard cases. Thus, these approaches still require a considerable amount of manual effort for fine-tuning and may not be applicable to SBDH factors with low prevalence. ^9^ Recent studies explored augmentation of low prevalence SBDH using simulated synthetic data and showed that fine-tuned Flan-T5 models outperformed zero-shot Generative Pretrained Transformer (GPT) models. ^16^

In this study, using the publicly available MIMIC-III dataset, ^17^ we analyzed all clinical notes for over 46,000 patients to identify 15 different SBDH categories using a well-known mathematical approach, called Latent Semantic Indexing (LSI). Using a subset of gold standard patient documents, we compared the performance of LSI with more recent GPT models.

## Methods

### Latent Semantic Indexing

The overview of our approach is shown in Figure 1.

**Figure 1:**
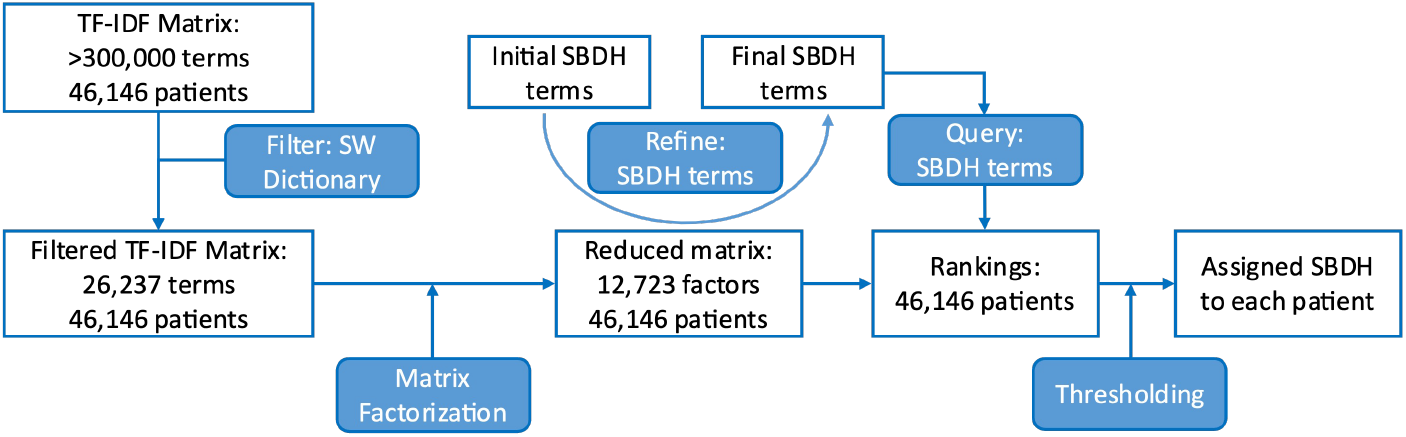
Workflow diagram of extracting and assigning SBDH factors to each patient in MIMIC-III dataset.

For each patient, a patient-document was created by concatenating the individual notes sequentially in the same order as present in the database. Terms (keywords) were extracted from patient documents using Text-to-Matrix Generator (TMG) package. ^18^ Punctuation (excluding hyphens and underscores) and capitalization were ignored. Additionally, articles and other common, non-distinguishing words were filtered out using the SMART stop list. ^19^ A term-by-patient matrix was created where the entries of the matrix were *tf-idf* weighted frequencies of terms across the patient document collection. Latent semantic indexing, a well-known factorization (Singular value decomposition) was performed on this matrix, subsequent to which each term and patient were represented as numeric vectors in reduced dimensions. The similarity between any two entities was calculated as the cosine between their respective vectors. The details of this process and various applications have been previously described by our group ^20–28^ and are documented in Additional file 1.

A total of 15 SBDH categories were considered, inspired from Social Determinants of Health (SDoH) categories defined by Torres et al., ^29^ and chronic behavior categories defined by the Center for Medicaid and Medicare Services (CMS). ^30^ The representative keyword for each category was finalized after consultation with a group of care managers. Table 1 lists the categories and their representative keywords while Supplementary Table S1 in Additional file 1 also lists the available ICD-9 codes for 9 of the 15 categories. For each keyword, patients were ranked in descending order of the cosine similarity between their truncated vectors. Patients with cosine scores *> Q*3 + (3.0 *∗ IQR*) were assigned to the respective SBDH category. The IQR (interquartile range) was calculated as Q3 (75^*th*^ percentile) – Q1 (25^*th*^ percentile).

**Table 1:**
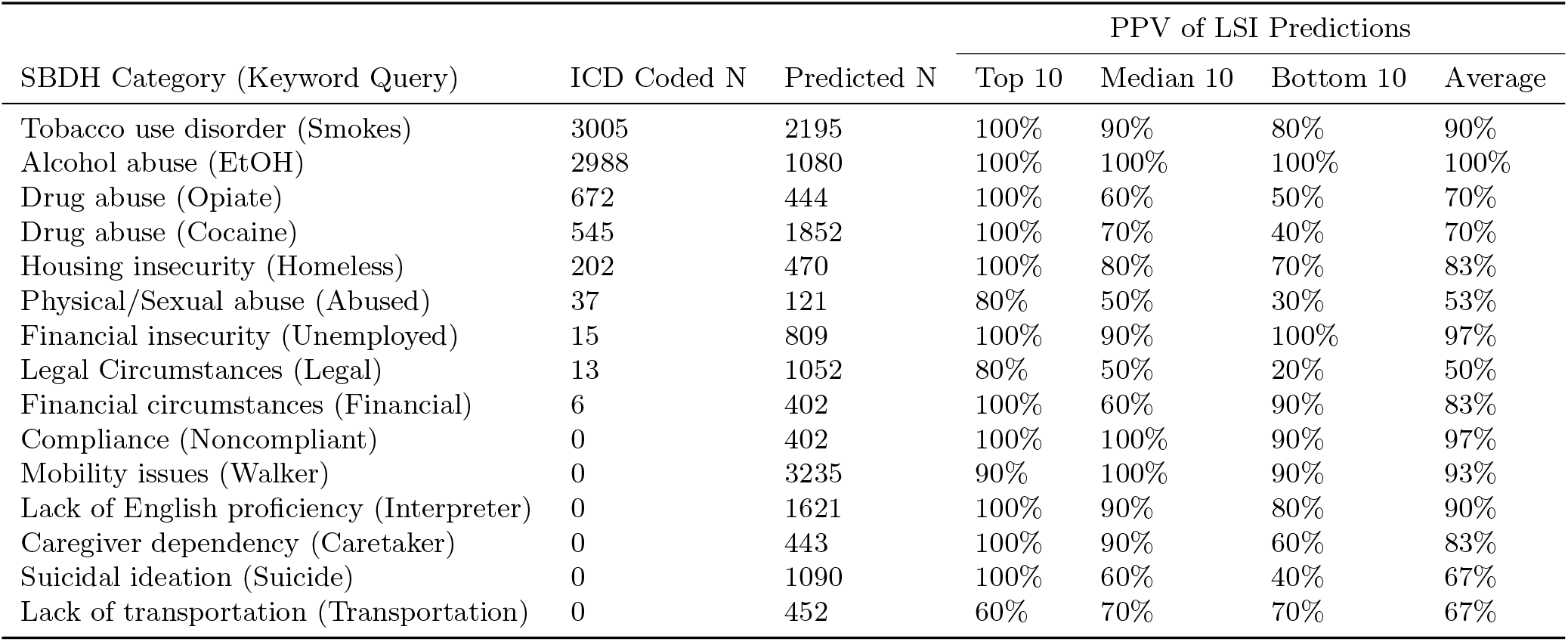
Performance of LSI predictions of SDBH categories. The terms in parentheses indicate the query word used to rank all patients in the dataset.

### Generative Pretrained Transformers (GPT)

All GPT API requests were made using a Python script which uses the “openai” library to connect to the OpenAI services in Azure, using gpt-3.5-turbo-1106 and gpt-4-1106-preview models. The Azure OpenAI

Service is a secure enterprise utility that is fully controlled by Microsoft and does not interact with any services operated by OpenAI (e.g. ChatGPT, or the OpenAI API). ^31^ Using this platform mitigated any potential risks to data sharing agreements or to patient privacy. Each API call included two components:

1) A function definition for the SBDH category, and 2) The contents of a patient document. GPT identifies the presence of the SBDH category in a document based on the name of the function and parameter names, with no other domain-specific information provided to the API. Each SBDH domain had its own function definition in the format of a JSON object (Additional file 1). Below is an example function definition for “Housing Insecurity”:

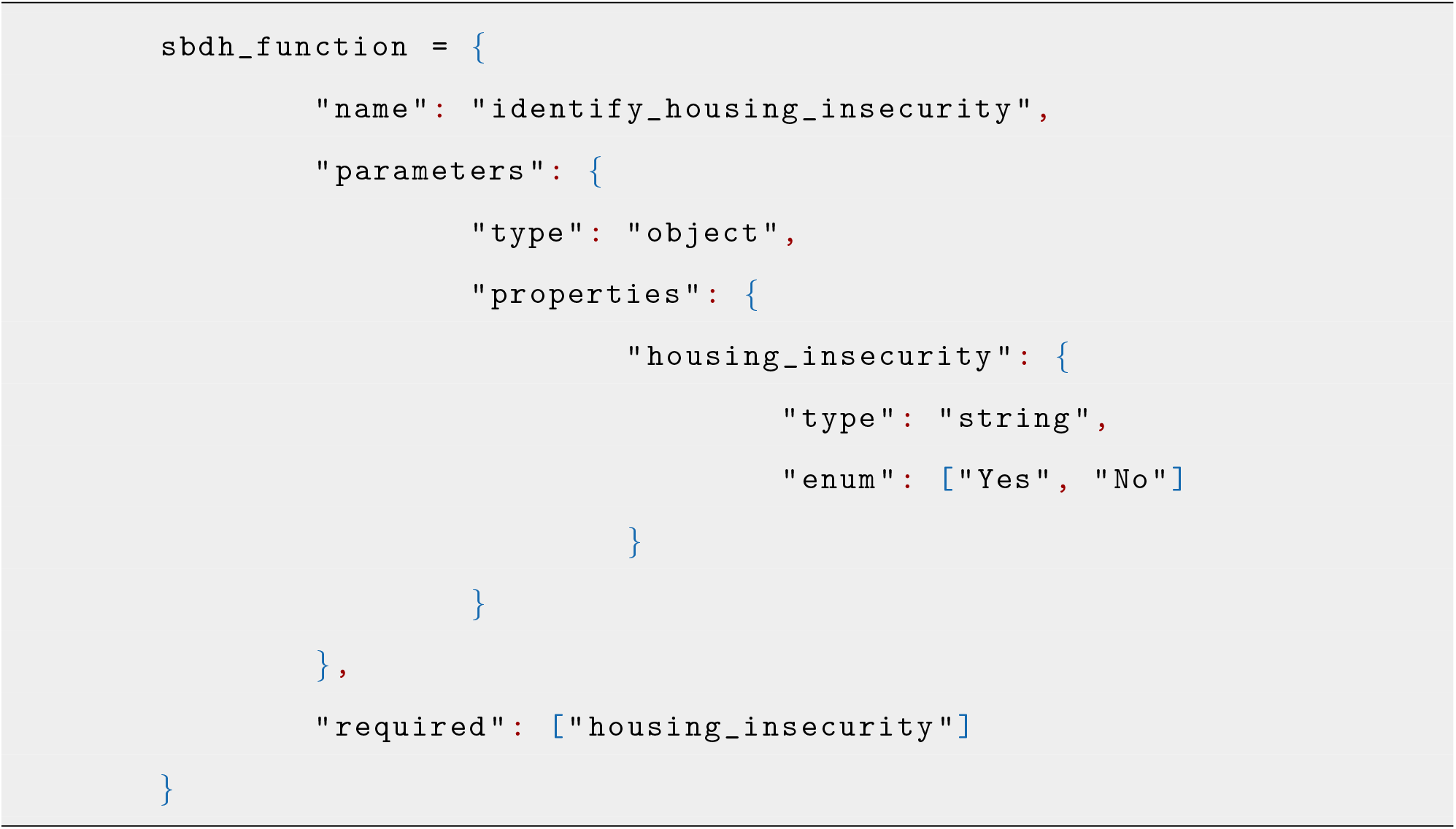

Sending a function ensures that the response from the API will be a predictable, well-formed JSON object with a binary answer of “Yes” or “No” to indicate the presence of the SBDH category in the patient document. The GPT engine does not actually call the function but instead treats the function like a callback, where the response from GPT includes the “Yes” or “No” value of the function parameter. The Python script calls the API as follows, including the patient document and the domain function as arguments:

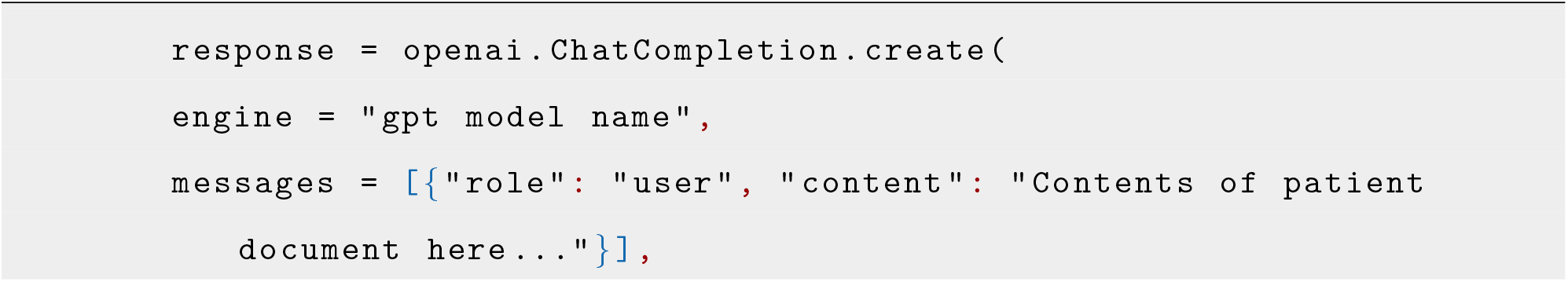
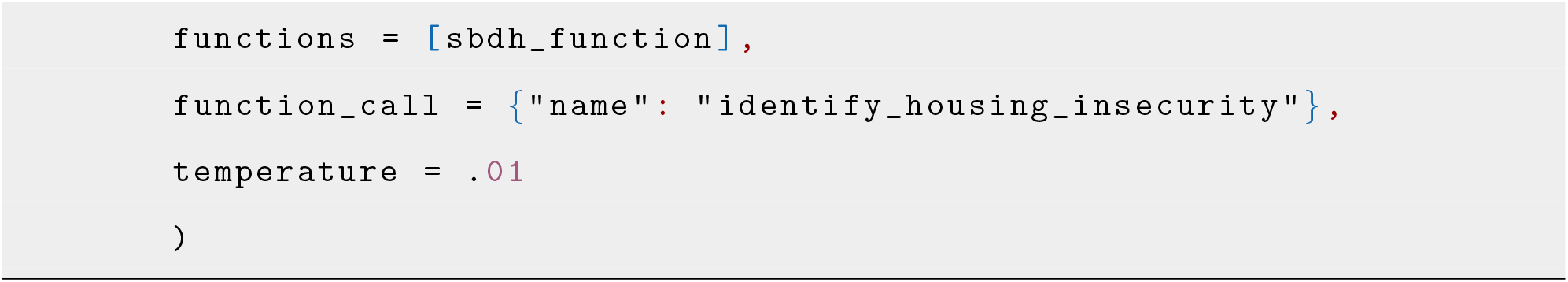

The “temperature” argument controls the determinism of the GPT model, accepting a value between 0 (more deterministic) and 2 (less deterministic). The API call and SBDH function definitions are identical for GPT-3.5 and GPT-4. All prompts were zero-shot, with no fine-tuning examples provided in the prompt.

### Analysis and Evaluation

The patient rankings pertaining to each SBDH keyword query was evaluated manually by chart review to determine the positive predictive value (PPV) of the top 10, median 10 and last 10 ranked. SBDH classification performance was evaluated using precision, recall and F1 score on manually curated gold standard (GS) samples. A random sample of up to 20 ICD-9 coded (when applicable) and up to 20 LSI-predicted cases were balanced with an equal number of non-coded and non LSI-predicted cases for each of the nine SBDH categories (that had at least six ICD-9 coded patients). This resulted in random samples ranging from 46 (financial circumstances) to a maximum of 80 (Tobacco use, Alcohol abuse and Opiate abuse). All cases were manually evaluated by chart review to determine actual positive (P) and negative (N) cases. Supplementary Table S3 in Additional file 1 includes the summary characteristics of the GS samples for each SBDH category. The performance of the text-based approaches was evaluated by Precision, Recall and F1 score.

To determine the overall performance of the text-based predictions using either LSI or GPT-4 in addition to ICD-9 coding, we used a logistic regression model including age, gender, race and ICD-9 for binary classification of GS patients corresponding to each SBDH category. In all three models, age was fit using a cubic spline with 2 degrees of freedom. The performance of each model was evaluated by 10-fold cross-validation and the Area Under the Receiver Operating Curve (AUROC).

## Results

A number of previous studies have demonstrated that International Disease Classification (ICD) codes cor-responding to social and behavioral determinants of health are not commonly used in the EHR. ^7^ Similarly, analysis of the MIMIC-III dataset showed that out of 44 potential Social Determinants of Health (SDoH) ICD-9 codes, ^29^ only 17 were used in MIMIC-III and only nine SDoH categories were assigned to three or more patients (Supplementary Figure S1 in Additional file 1). To develop a comprehensive set of SBDH for benchmarking the text-based approaches, we included the following SDoH categories in order of frequency: *Lack of housing* (202), *history of physical abuse* (37), *unemployment* (15), *legal circumstances* (13), *inadequate material resources* (6). In addition, we included four behavioral chronic conditions defined by CMS ^30^ and several other SBDH categories such as suicide ideation and compliance, which are represented in ICD-10 but not in ICD-9. Altogether, this study focused on 15 SBDH categories (Table 1), although only nine categories were documented by ICD-9 billing codes in this data set (Supplementary Table S1 in Additional file 1).

### Latent Semantic Indexing and Lexicon Development

Latent Semantic Indexing (LSI) is a well-known matrix factorization method, which reduces the dimensionality of terms and documents in to lower rank matrices. ^20–28^ By using a lower rank matrix, the terms can be grouped together more conceptually, whereas by using higher ranks, terms can be grouped more literally. In addition, patients can be grouped together in more conceptual or literal fashion based on the content in their clinical notes.

Out of a total of 46,520 patients in the MIMIC-III dataset, 46,146 patients had clinical notes. The number of notes associated with these patients ranged from 1 to 1420, with the median being 21 notes. A patient document was constructed by concatenating all clinical notes together for each patient, which resulted in a term dictionary of *>*300,000 terms. To reduce the dictionary size to terms that are relevant to SBDH, we filtered the dictionary to include only terms that were extracted from social history sections, resulting in a final dictionary size of 26,237 terms. Each term in the 26,237 terms-by-46,146 patients matrix was weighted using *tf-idf* and then factorized to 12,723 dimensions (see Additional file 1 for details).

To determine the best lexicons (terms) to represent various SBDH categories, we manually constructed a set of 134 keywords (including variants, plurals and common misspellings) corresponding to the SBDH categories described above (Supplementary Table S2 in Additional file 1). Both the SBDH categories and the lexicons were iteratively refined as described below based on: 1) The correlations between terms with respect to the vector of all ranked patients in the MIMIC-III dataset (Figure 2a), 2) the precision of the top ranked patients for the keyword query, 3) the recall of ICD-9 coded patients.

**Figure 2:**
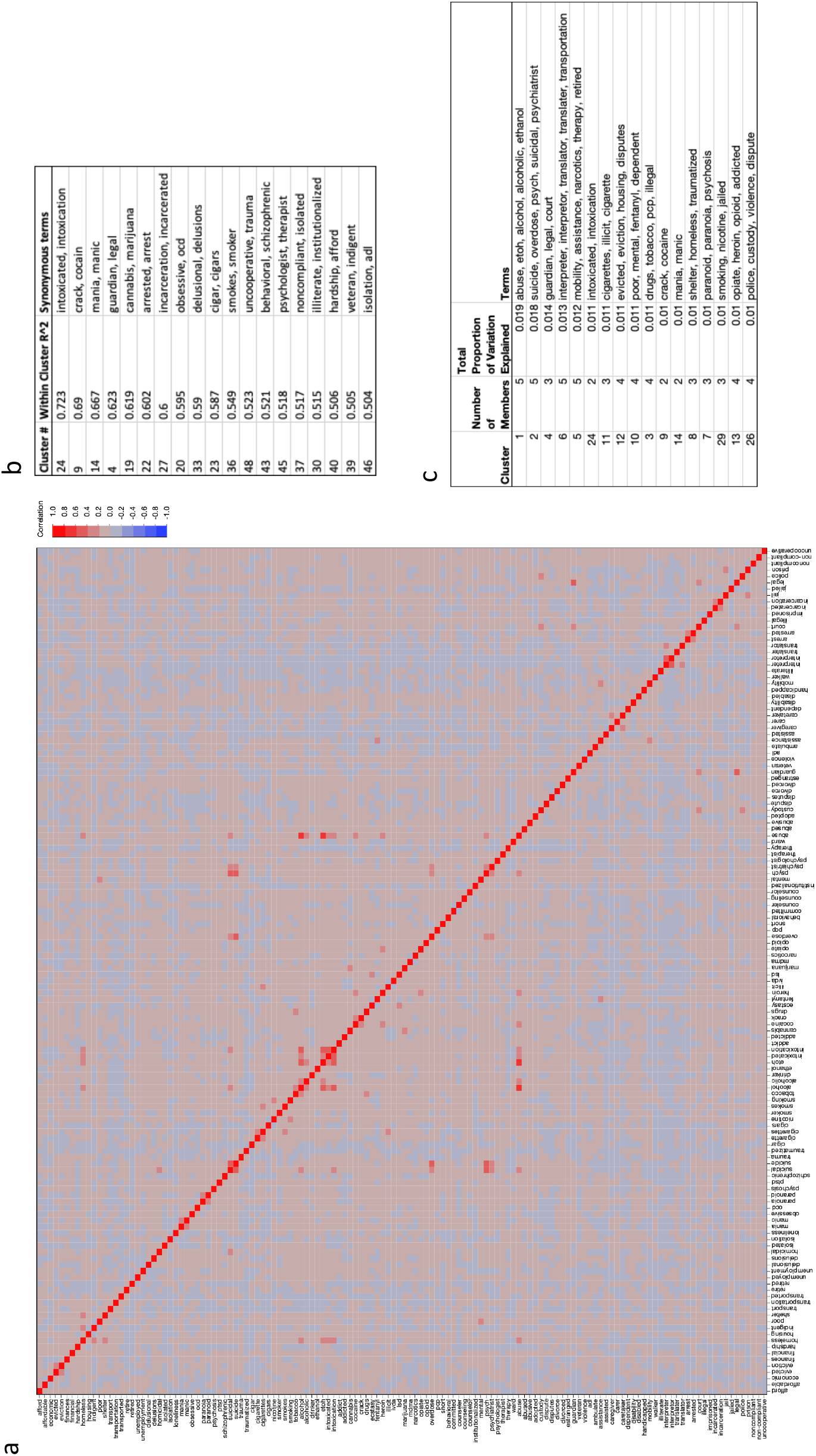
Relationship between SBDH terms in reduced-rank (12,723) vector space model. a) Heatmap of correlations between terms, where red represent high correlation and blue represents low correlation. b) List of clusters with the highest intra-cluster correlations, depicting terms that are explicitly or conceptually synonymous as well as terms that share stems. c) List of terms in clusters that account for 20% of the variability in the entire patient population.

Clustering of the term correlations revealed groups of highly synonymous terms deduced from the word usage patterns in the patient documents. This demonstrates the utility of matrix factorization as an unsupervised machine learning approach which learns conceptually related terms based on the word usage patterns in the clinical notes. For example, factorization revealed that words such as intoxicated/intoxication, crack/cocaine, or manic/mania are synonymously used in the clinical notes (Figure 2b). In addition, this approach identified short phrases in a rudimentary way, such as legal/guardian (Figure 2b). Lastly, some of the larger clusters included broader contextual information, such as suicide/overdose/psych/suicidal/psychiatrist (Figure 2c).

### Evaluation of LSI-derived SBDH Predictions

All patients in the collection were ranked based on a representative keyword query for each of the 15 SBDH categories. Application of interquartile outlier detection method determined the cosine threshold for each query where the patients ranked above the threshold (*> Q*3 + (3.0 *∗ IQR*)) are highly associated with the query and thus predicted to have the specific SBDH. In all but three SBDH categories (Tobacco use, Alcohol abuse, and Drug abuse -Opiate), the number of patients in the collection with an LSI-predicted SBDH were substantially higher than the ICD-9 coded patients (Table 1).

To evaluate the classification performance of the SBDH predictions, we determined the PPV by manual evaluation of the top 10, median 10, and bottom 10 patients within the cut-off threshold (Table 1). In all but four SBDH categories, the PPV of the top 10 ranked patients was 100%. As expected, the PPV decreased with lower rankings. The average PPV for all 15 SBDH categories ranged from 50% (legal circumstances) to 100% (alcohol abuse), with nine of the SBDH categories having a PPV *≥* 83%.

Next, we compared the performance of ICD-9 coding with either LSI, GPT-3.5 or GPT-4 large language models using different sets of gold standard (GS) patients that were randomly selected for each SBDH category and manually labeled by chart review. The characteristics of the GS sets of patients for each SBDH category are provided in Supplementary Table S3 in Additional file 1. Only nine SBDH categories that had at least six ICD-9 coded patients were included in this analysis. Importantly, only LSI was able to process all of the patient documents. In contrast, due to context window size restrictions, GPT-3.5 processed 55.6% of the gold standard documents and GPT-4 processed 94.2% (Figure 3).

**Figure 3:**
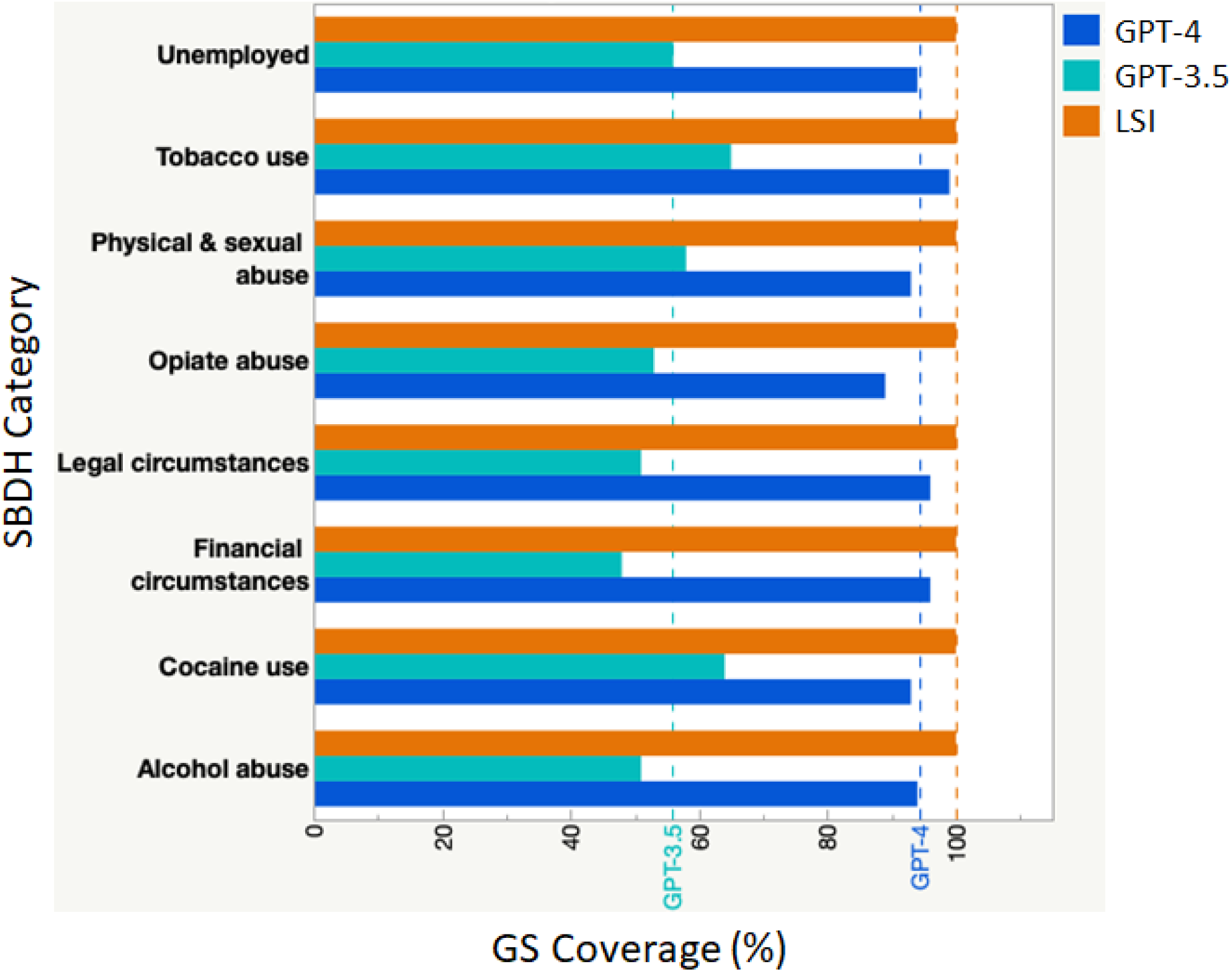
Proportion of gold standard patient documents for each SBDH category that yielded results by LSI, GPT-3.5 or GPT-4.0.

Earlier versions of GPT were highly irreproducible such that the same prompt could produce different responses or no response at all. To evaluate this phenomenon, we compared the responsiveness of GPT-3.5 and GPT-4 to the same set of shared documents within the 16K context window limit of GPT-3.5 for each of the nine SBDH categories (Table 2). For GPT-3.5, the same set of documents were submitted using the same prompt five independent times. GPT-3.5 was unresponsive for 2% (Cocaine use) to 30% (unemployed) of the patient documents across the SBDH categories. In addition, in all but one SBDH category, GPT-3.5 provided conflicting responses between the five independent prompts. For example, although GPT3.5 provided responses for all 27 patient documents related to legal circumstances, it provided conflicting responses for six (22%) of the patient documents (Table 2). In contrast, GPT-4 was unresponsive for only two documents (3.8%) in only one SBDH category (tobacco use).

**Table 2:** Unresponsiveness of GPT-3.5 and GPT-4. On a set of shared patient documents (N), GPT-3.5 was prompted five independent times, whereas GPT-4 was prompted only once. The % of documents where GPT-3.5 or GPT-4 did not provide a response is iindicated for each SBDH category. The % disagreement corresponds to the number of documents where GPT-3.5 provided conflicting binary responses.

| SBDH Category | N | GPT-3.5 |  | GPT-4 |
| --- | --- | --- | --- | --- |
|  |  | % Disagreement | % No Response | % No Response |
| Housing insecurity | 48 | 6.3% | 0.0% | 0.0% |
| Tobacco use | 52 | 3.8% | 15.4% | 3.8% |
| Opiate abuse | 42 | 7.1% | 0.0% | 0.0% |
| Alcohol abuse | 41 | 2.4% | 0.0% | 0.0% |
| Cocaine use | 51 | 0.0% | 2.0% | 0.0% |
| Physical & sexual abuse | 39 | 2.6% | 5.1% | 0.0% |
| Unemployed | 30 | 6.7% | 30.0% | 0.0% |
| Legal circumstances | 27 | 22.2% | 0.0% | 0.0% |
| Financial circumstances | 22 | 13.6% | 4.5% | 0.0% |

As expected, due to the limitations described above, the average recall of GPT-3.5 across all of the documents in all nine SBDH categories was low (0.41), compared to LSI (0.70) and GPT-4 (0.77) (Table 3). Overall, the average macro-F1 was highest for GPT-4 (0.8), followed by LSI (0.74), ICD-9 (0.71) and GPT-3.5 (0.54) despite the fact that GPT-4 was unable to process 5.8% of the documents due to context window size limitations (Figure 3 & Table 3).

**Table 3:**
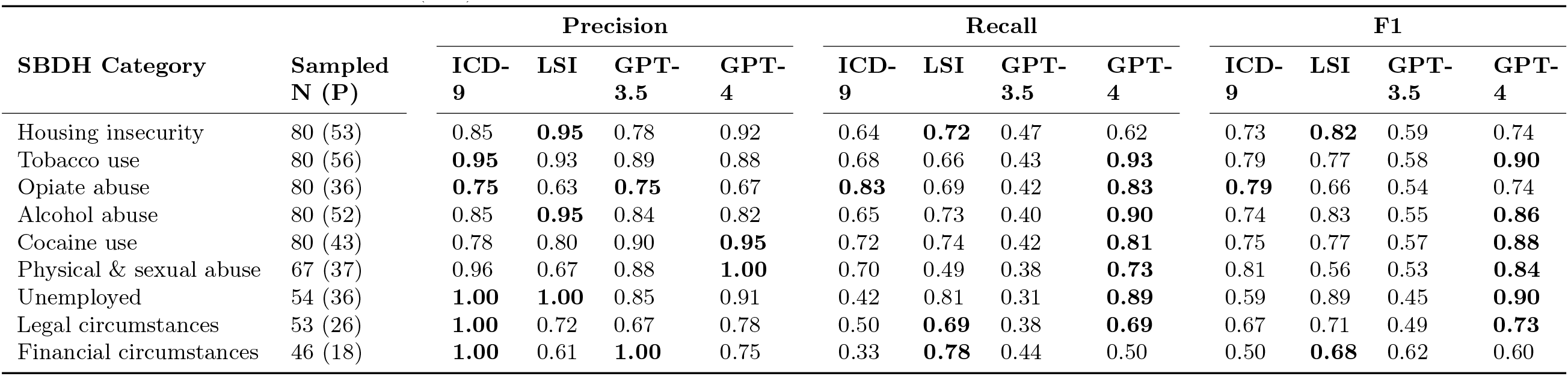
Retrieval performance of each method alone using a set of sampled Gold Standard cases. The bold text indicate the highest precision, recall, or F1 for each SBDH category (row).

It is important to note that in some cases, although a patient was assigned an ICD-9 code for a particular SBDH, supporting documentation in the clinical notes could not be found. In such cases, the ICD-9 coded individuals were assumed to be actual positives. Therefore, to retrieve all possible SBDH in a given GS set, the text-based prediction of SBDH was combined with ICD coded individuals across the nine SBDH categories. On average, all three methods performed similarly with respect to precision, recall and F1 when combined with ICD-9 (Figure 4).

**Figure 4:**
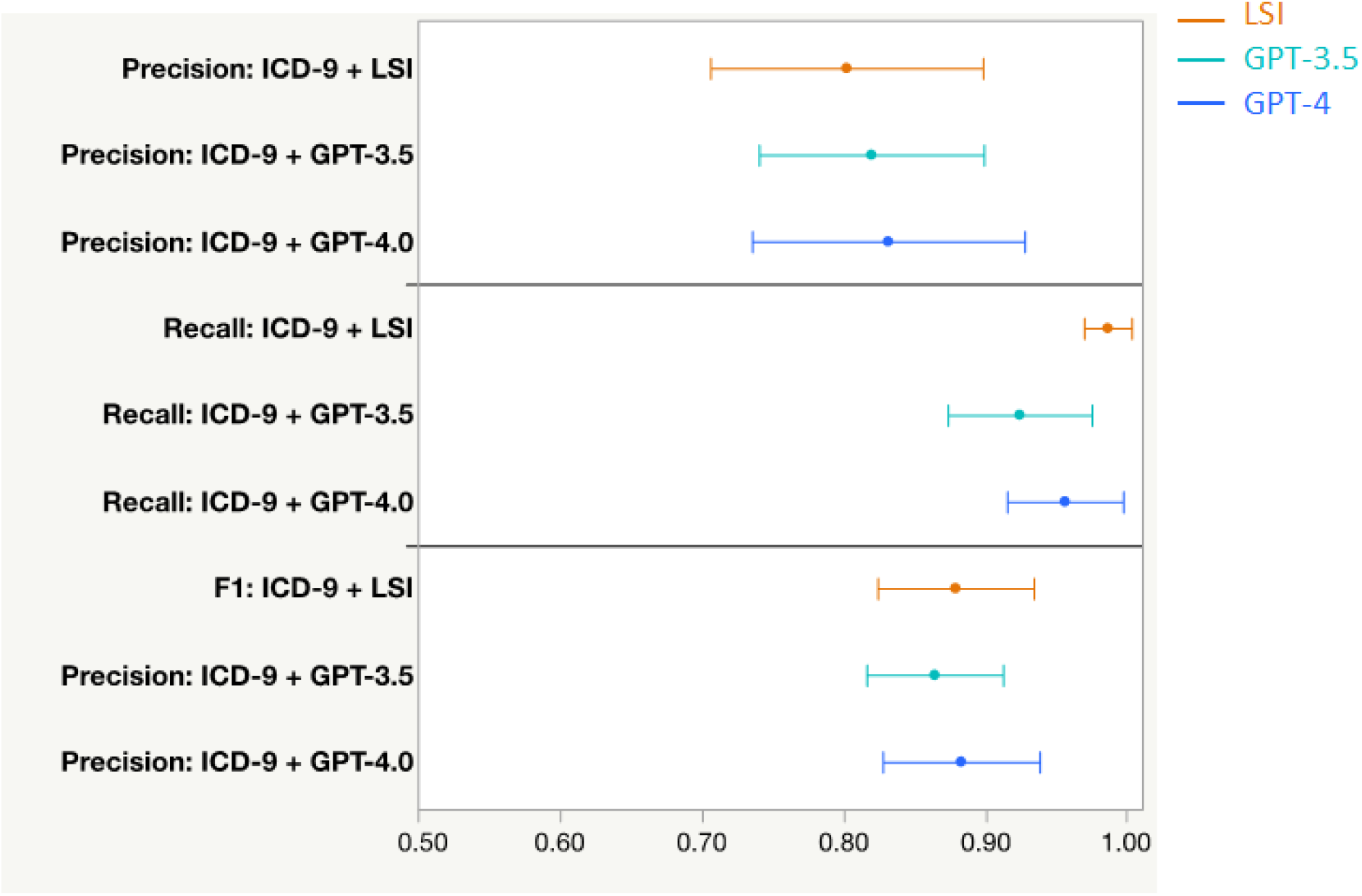
Retrieval performance of LSI, GPT-3.5 or GPT-4 when combined with ICD-9 coding. Precision (upper panel), recall (middle panel) and F1 (lower panel) of ICD-9 combined with either LSI (orange lines), GPT-3.5 (cyan lines) and GPT-4 (blue lines). Values represent the mean (filled circle) and 95% confidence intervals (error bars) across the nine SBDH gold standard sets.

Lastly, to evaluate the overall predictive performance of LSI with GPT-4 when combined with ICD-9 coding, we compared the prediction AUC of three different logistic regression models: 1) base model including gender, age, race and SBDH ICD-9 codes, 2) base model plus LSI identified SBDH, 3) base model plus GPT-4 identified SBDH (Figure 5). Using only ICD-9 coding (base model), the AUCs for the nine SBDH categories ranged between 0.69 (*housing insecurity* and *financial circumstances*) to 0.85 (*history of physical and sexual abuse*). In all nine categories, inclusion of LSI or GPT-4 improved the AUCs compared to ICD-9. Interestingly, LSI outperformed GPT-4 in six of the nine SBDH categories (*housing insecurity, unemployment, opiate abuse, alcohol abuse, legal circumstances*, and *financial circumstances*).

**Figure 5:**
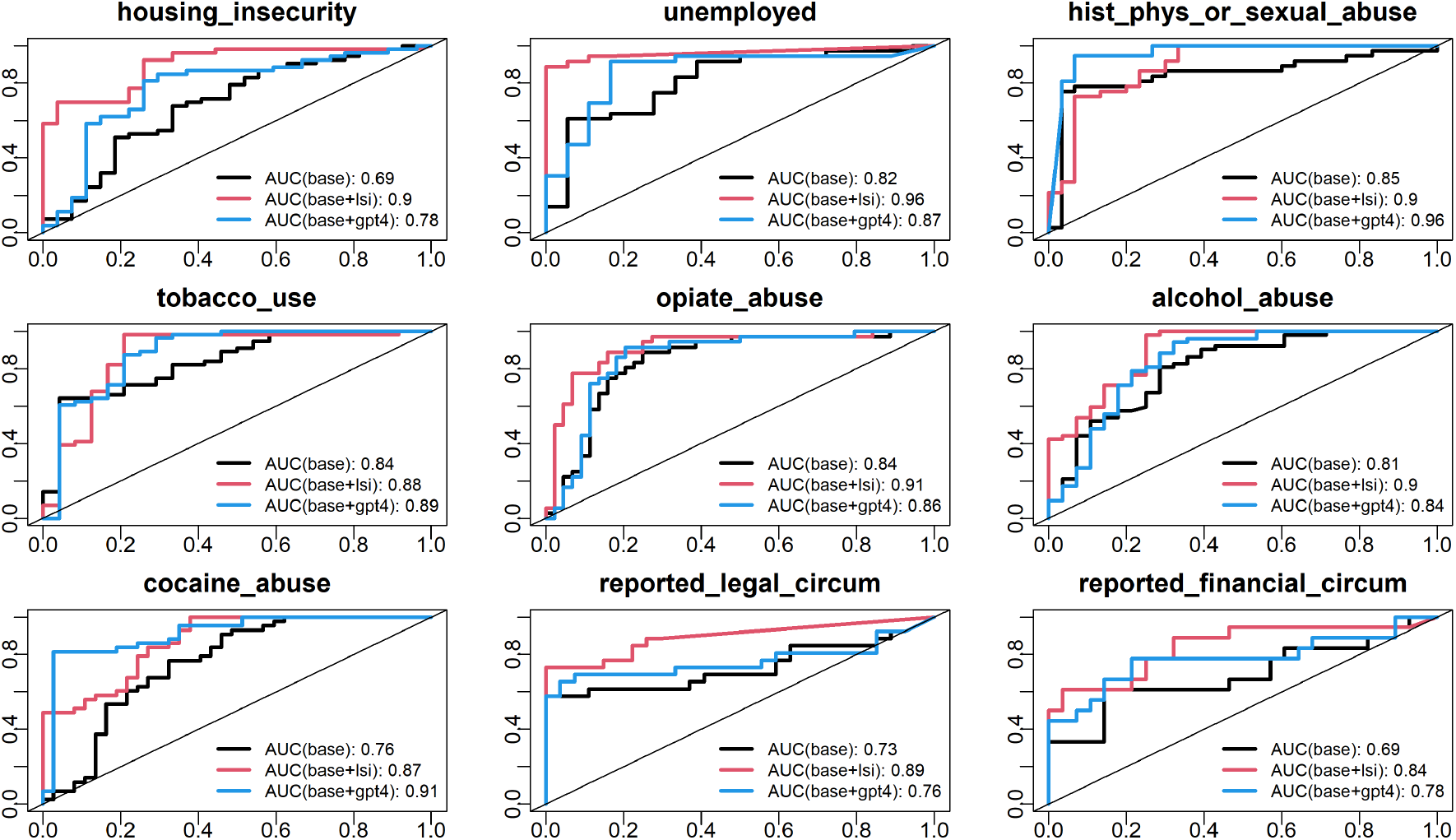
Comparison of classification performance of ICD-9 and/or text-predicted SBDH cat-egories using multivariable analysis. The AUC is shown for three different models: 1) Base model including age, gender and ICD-9 codes (black lines), 2) Base model plus LSI identified SBDH (red lines), and 3) Base model plus GPT-4 identified SBDH (blue lines).

## Discussion

In this study, we demonstrated the utility of LSI as a robust unsupervised approach for comprehensively processing all clinical notes in the EHR to identify SBDH and to supplement the SBDH documented by ICD-9 diagnosis codes. Importantly, we show that although LSI is a bag-of-words approach, it performed similarly and sometimes better than GPT models. This work highlights several advantages for using LSI in real-world healthcare applications.

One major advantage of LSI is its ability to process all of the notes for a given patient without the imposed context window token size limitations of GPT. As pointed out in Figure 3, only 55.6% and 94.2% of the GS cases could be processed by GPT-3.5 and GPT-4, respectively. At the time of our analysis, the input context window size limits for GPT-3.5 and GPT-4 were 16K and 128K tokens, respectively. However, other LLMs may have larger context windows. Even with the context window limits, it is possible to process larger documents by ‘chunking’, a method where a large document is split into smaller overlapping documents that are smaller than the token limits. In our analysis, we did not attempt to process all of the GS documents, instead we directly compared the performance of LSI with GPT-3.5 and GPT-4 using the same set of documents (Table 3 and Figures 4 & 5). Another reason for limiting the analysis to a subset of GS documents was cost. At the time of the analysis, the cost for GPT-3.5 and GPT-4 using the Microsoft Azure OpenAI ^31^ services per query was USD $0.001 and $0.01 per 1K input tokens, respectively. Thus, it would have been more costly to chunk the larger GS documents. Another way to reduce the number of GPT queries would have been to perform multi-class labeling. In our analysis, we performed single class labeling, where each document was processed individually to identify a single SBDH category. Although this approach would be useful, it may require considerable fine-tuning and may not be feasible for identifying all 15 SBDH categories at once.

Another major advantage of LSI is that it does not require external training on a large dataset and fine-tuning for domain specific applications. For this study, the LSI model was built using all of the clinical notes for all of the *>* 46, 000 patients at once. In contrast, GPT and other LLM require extensive training using large amounts of external data sources. For example, GPT 3.5 was trained on 175 billion parameters using training data up to September 2021. Although the models perform well for general text analysis, they may not perform well on specialized clinical tasks. For example, Lybarger et al. developed an event based deep-learning extractor for SBDH that determines chronicity, duration, frequency and type of event. ^12^ However, their models apply only to a subset of SBDH categories, including employment, living status, as well as alcohol, tobacco and drug use. They point out that training these models required significant manual effort by human experts to develop both positive and negative gold standard datasets for fine-tuning. ^12^ In addition, since these methods require large amounts of training data for fine-tuning, they can have limited usefulness for SBDH categories that are rare (low prevalence).

Another major advantage of LSI is that, unlike GPT, it is deterministic (reproducible) and 100% responsive to all queries. For a given factorization rank, LSI produces the same exact ranking of the documents based on the same query. On the other hand, we showed (Table 2) that GPT-3.5 produces conflicting responses to the same prompt on the same set of documents. Moreover, we demonstrated that both GPT-3.5 and GPT-4 may not respond, a phenomenon commonly referred to as ‘laziness’. Although the GPT-4 model has been improved to reduce laziness, we found that it can be unresponsive as the document size reaches its maximum context window size limits.

Our findings indicate that using clinical notes to identify SBDH should not replace efforts in health systems to screen for SBDH, rather provide a complementary approach to enhance estimates of the SBDH burden (prevalence) in large populations. During chart review for developing the GS sets, we found a few ICD-9 coded individuals who had no supporting documentation for the codes. For example, some patients had few encounters with the health system and had no social history notes, yet were coded for homelessness or alcohol abuse. As reported by others, this observation illustrates the importance of combining the information provided by ICD-9 codes and other structured data (e.g., questionnaires) with unstructured data in the EHR to obtain a more representative assessment of the SBDH prevalence in a population. ^7,10–12,32^ On the other hand, implementing SDoH screening tools across a large health system is impractical and potentially biased. Studies have shown that SDoH screening forms are primarily implemented in outpatient and primary care settings. However, it is thought that socioeconomically disadvantaged individuals are less likely to go to primary care, instead use the emergency department (ED) for their healthcare needs. ^33^ Moreover, a recent study demonstrated that only 3.7% of the patients in a large health care system in South Carolina had answered all 11 questions on the SDoH screening forms. ^34^ Therefore, for better assessment of SBDH burden in a population, information must be aggregated from a variety of sources in the EHR, including the clinical notes.

It is worth highlighting that the costs associated with OpenAI services make it currently unrealistic to implement in health systems to assess SBDH burden in large populations of patients. To address this issue, future research will focus on using LSI to narrow large populations of patients into smaller groups that are conceptually predicted to have SBDH and then process those documents using GPT to contextualize and validate the LSI predictions. Factorization provides value beyond keyword searching alone because it contextualizes keywords as vectors in reduced ranked space, thereby grouping words that are frequently used together in the context of SBDH keywords. This approach provides a general advantage by automatically grouping synonyms, misspellings, and conceptually related terms that are often used together in narratives (Figure 2). For example, a homeless individual is often unemployed and has drug/alcohol abuse problems. Also, factorization is able to infer that ‘shelter’ and ‘homelessness’ are synonymously used in the narratives. By lowering the rank of the factorized matrix, one can identify a subset of patients who are conceptually related to the SBDH, achieving higher recall than precision. By subsequently processing these patient documents with GPT-4, the specific evidence in support of the SBDH can be readily deduced while keeping the overall processing cost low.

While LSI was highly sensitive (high PPV) for most SBDH categories, its performance was limited for a few SBDH categories such as legal circumstances. We found that legal circumstances covered a broad range of areas ranging from power of attorney, guardianship issues, hospital liability to encounters with law enforcement for illegal activities. More refinement would be necessary to evaluate the performance of our approach on specific areas pertaining to specific legal circumstances. For example, guardianship issues for clinical decision making could be better identified with a ‘guardian’ query rather than a general term such as ‘legal’. In three cases (alcohol abuse, tobacco use, and opiate abuse), our approach identified fewer cases than ICD coded individuals. This may be due to the fact that drug, alcohol and tobacco use are routinely captured within structured fields in current clinical practice. However, other SBDH categories are not routinely captured. One approach to increase the number of cases identified by our approach would be to relax the thresholding parameter or to combine multiple lexicons representing alcohol abuse in an additive way.

Feller et al. were among the first groups to apply NLP methods to infer SBDH from clinical notes. After feature selection, they included 2-4,000 individual words as independent variables in various machine learning classifiers to identify sexual history, sexual orientation, alcohol use, substance use and housing status. They found that combining clinical notes and structured data enabled reasonably accurate inference of these SBDH categories. ^35,36^ Bejan et al., using a vector embedding approach to expand SDoH lexicons, demonstrated better performance of identification of homelessness and adverse childhood experiences (ACEs) from clinical notes. ^37^ Our process, which combines the bag-of-words approach with factorization for embedding, allows an automated method to identify a broad set of SBDH categories.

The LSI approach has several limitations. First, it is a bag-of-words embedding technique, which does not account for word context (phrases) and negated terms. In addition, the performance of our approach was affected by the presence of forms and templated text in the clinical notes, such as “Family Information” or social history forms, where there are many negations and repeated text. The performance of our approach would improve if certain note types, forms and templates were removed during pre-processing. Lastly, our approach does not provide temporal relations and event-types. As stated above, many of these limitations would be addressed by combining the advantages of LSI (e.g., robustness, determinism, and no cost) with the advantages of LLM (i.e., contextualization, removal of negation, and multi-label classification).

## Conclusions

In this study, we demonstrated that using an unsupervised machine learning factorization approach on clinical notes is a robust way to enhance SBDH identification from the EHR. This work is significant because it provides an automated way to extract SBDH for patients in a health system without the additional burden of implementing standardized surveys in clinical workflows. By providing better estimates of SBDH burden in populations, this work sets the stage for developing patient level health risk and utilization prediction models that incorporate SBDH factors in addition to standard clinical and structured data from the EHR.

## Data Availability

The MIMIC-III dataset is available publicly through physionet.org.

## Declarations

### Ethics approval and consent to participate

Not applicable.

## Consent for publication

Not applicable.

## Availability of data and materials

The MIMIC-III dataset is available publicly through physionet.org.

## Authors’ contributions

SR designed and implemented the methods, generated data, interpreted results and contributed to writing of the manuscript. SM generated data and performed analysis. LZ analyzed data, interpreted results and contributed to writing of the manuscript. RH designed the study, interpreted results, performed chart reviews and wrote the manuscript.

## Acknowledgements

The authors are grateful to Oakland University for providing the high-performance computing resources and to MIT Laboratory for Computational Physiology for providing the MIMIC-III dataset. We thank Kevin Heinrich (Quire Inc.) and Brad Silver (Quire Inc.) for helpful discussions.

