## Additional file 1 for "Large-scale identification of social and behavioral determinants of health from clinical notes: Comparison of Latent Semantic Indexing and Generative Pretrained Transformer (GPT) models"

Social determinants of health, electronic health records, machine learning, natural language processing, clinical notes

#### SUPPLEMENTARY FIGURES and TABLES

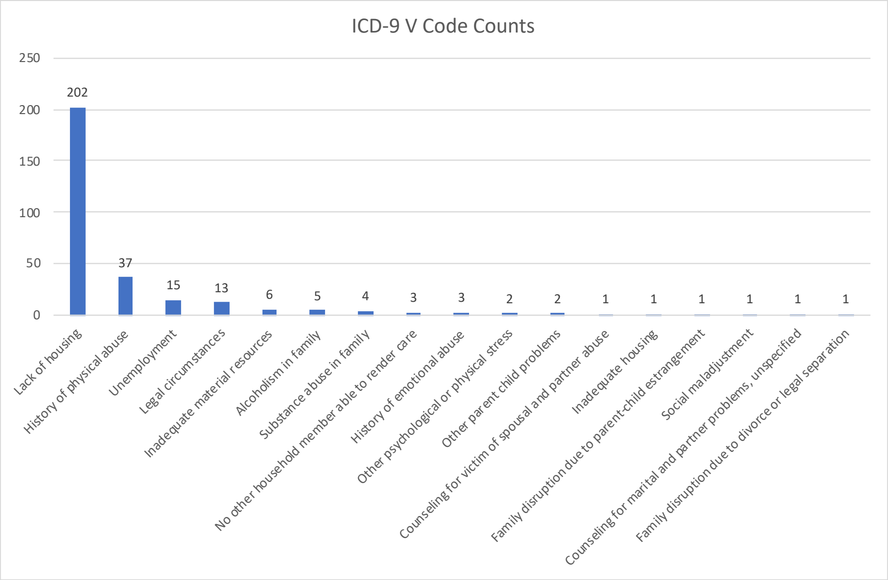


Figure S1: Distribution of patient counts with SDoH categories.

Table S1: ICD-9 codes for SBDH categories.

| SBDH Category | ICD-9 Code | ICD9 Description | Keyword |
|---|---|---|---|
| Housing insecurity | V600 | Lack of housing | Homeless |
| Physical/Sexual abuse | V1541 | History of physical abuse | Abused |
| Financial insecurity | V620 | Unemployment | Unemployed |
| Legal circumstances | V625 | Legal circumstances | Legal |
| Financial circumstances | V602 | Inadequate material resources | Financial |
| Tobacco use | 3051 | Tobacco use disorder | Smokes |
| | 64903 | Tobacco use disorder complicating pregnancy, childbirth, or the puerperium, antepartum condition or complication | |
| Alcohol abuse | 30500 | Alcohol abuse, unspecified | Etoh |
| | 29181 | Alcohol withdrawal | |
| | 30391 | Other and unspecified alcohol dependence, continuous | |
| | 30390 | Other and unspecified alcohol dependence, unspecified | |
| | 30501 | Nondependent alcohol abuse | |
| | 30301 | Acute alcoholic intoxication in alcoholism, continuous | |
| | 2910 | Alcohol withdrawal delirium - as a primary diagnosis code, | |
| | 5711 | Acute alcoholic hepatitis - as a primary diagnosis code, | |
| | 30300 | Acute alcoholic intoxication-unspec | |
| | 4255 | Alcoholic cardiomyopathy | |
| | 9800 | Toxic effect of ethyl alcohol | |
| | 5710 | Alcoholic fatty liver | |
| | 3575 | Alcoholic polyneuropathy | |
| | 30502 | Alcohol abuse, episodic | |
| | 30392 | Other and unspecified alcohol dependence | |
| | 5713 | Alcoholic liver damage, unspec. | |
| | 2911 | Alcohol-induced persisting amnestic disorder | |
| | 53530 | Alcoholic gastritis, without mention of hemorrhage | |
| | 2912 | Alcohol-induced persisting dementia | |
| | E8600 | Acc poisn-alcohol bevrag | |
| | 2913 | Alcohol-induced psychotic disorder with hallucinations | |
| | 53531 | Alcoholic gastritis, with hemorrhage | |
| | 30302 | Acute alcoholic intoxication. | |
| Drug abuse - Cocaine | 30560 | Cocaine abuse-unspec | Cocaine |
| | 30561 | Cocaine abuse, uncomplicated | |
| | 30421 | Cocaine dependence, continuous | |
| | 30420 | Cocaine dependence, unspecified | |
| | 30562 | Cocaine abuse-episodic | |
| | 76075 | Mat cocaine aff NB/fet. | |
| | 30422 | Cocaine dependence, episodic | |
| Drug abuse - Opiate | 96509 | Poisoning by other opiates and related narcotics | Opiate |
| | E8502 | Accidental poisoning by other opiates and related narcotics | |
| | 30401 | Opioid type dependence, continuous | |
| | 30550 | Opioid abuse, unspecified | |
| | 30400 | Opioid type dependence, unspecified | |
| | 30551 | Opioid abuse, continuous | |
| | 30470 | Combinations of opioid type drug with any other drug dependence, unspecified | |
| | 30471 | Combinations of opioid type drug with any other drug dependence, continuous | |
| | 30552 | Opioid abuse, episodic | |
| | 30402 | Opioid type dependence, episodic | |

Table S2: SBDH categories and their associated keywords (134) that were manually selected. The keywords that were chosen as the best representative of the respective SBDH categories are italicized.

| **SBDH Categories (Refined Categories)** | **Keywords** |
|---|---|
| Alcohol Use Disorders (Alcohol Abuse) | alcohol, alcoholic, drinker, ethanol, *etoh*, intoxicated, intoxication |
| Care Provider Dependency (Caregiver dependency, Mobility issues) | adl, ambulate, assistance, assisted, caregiver, carer, *caretaker*, dependent, disability, disabled, handicapped, mobility, *walker* |
| Compliance | *noncompliant*, non-compliant, uncooperative |
| Drug Use Disorders (Drug abuse - Cocaine, Drug abuse - Opiate) | addict, addicted, cannabis, *cocaine*, crack, drugs, ecstasy, fentanyl, heroin, illicit, ivda, lsd, marijuana, mdma, narcotics, *opiate*, opioid, overdose, pcp, snort |
| Education and Literacy (Lack of English Proficiency) | illiterate, *interpreter*, interpretor, translater, translator |
| Family Circumstances (Physical/Sexual Abuse) | abuse, *abused*, abusive, violence, adopted, custody, dispute, disputes, divorce, divorced, estranged, guardian, veteran |
| Housing & Economic Circumstances (Financial Circumstances, Housing Insecurity, Lack of Transportation) | afford, affordable, economic, evicted, eviction, finances, *financial*, hardship, *homeless*, housing, indigent, poor, retire, retired, shelter, transport, *transportation*, transported |
| Legal Circumstances | arrest, arrested, court, illegal, imprisoned, incarcerated, incarceration, jail, jailed, *legal*, police, prison |
| Other Psychosocial Circumstances (Suicide Ideation) | behavioral, committed, counseler, counseling, counselor, delusional, delusions, homicidal, institutionalized, isolated, isolation, loneliness, mania, manic, mental, obsessive, ocd, paranoia, paranoid, psych, psychiatrist, psychologist, psychosis, ptsd, schizophrenic, suicidal, *suicide*, therapist, therapy, trauma, traumatized, ward |
| Tobacco use | cigar, cigarette, cigarettes, cigars, nicotine, smoker, *smokes*, smoking, tobacco |
| Unemployment (Financial Insecurity) | *unemployed*, unemployment |

Table S3: Characteristics of independently sampled gold standard (GS) patients for each SBDH category.

| Variable | Alcohol Abuse | Drug Abuse, Cocaine | Drug Abuse, Opiate | Financial Circumstances |
|---|---|---|---|---|
| n | 80 | 80 | 80 | 46 |
| Male, n (%) | 62 (77.5%) | 52 (65.0%) | 52 (65.0%) | 29 (63.0%) |
| Age, Median (Q1, Q3) | 51.0 (40.0, 60.0) | 44.0 (33.5, 52.0) | 47.0 (32.0, 57.5) | 57.0 (42.0, 69.0) |
| Ethnicity, n (%) | | | | |
| - ASIAN | 1 (1.2%) | 0 (0.0%) | 3 (3.8%) | 2 (4.3%) |
| - BLACK/AA | 5 (6.2%) | 6 (7.5%) | 4 (5.0%) | 6 (13.0%) |
| - HISPANIC OR LATINO | 4 (5.0%) | 9 (11.2%) | 1 (1.2%) | 3 (6.5%) |
| - OTHER | 10 (12.5%) | 13 (16.2%) | 18 (22.5%) | 7 (15.2%) |
| - WHITE | 60 (75.0%) | 52 (65.0%) | 54 (67.5%) | 28 (60.9%) |
| Text, n (%) | 40 (50.0%) | 40 (50.0%) | 40 (50.0%) | 23 (50.0%) |
| ICD-9, n (%) | 40 (50.0%) | 40 (50.0%) | 40 (50.0%) | 6 (13.0%) |
| True Positive, n (%) | 52 (65.0%) | 43 (53.8%) | 36 (45.0%) | 18 (39.1%) |

| Variable | Financial Insecurity | Housing Insecurity | Legal Circumstances | Physical/Sexual Abuse |
|---|---|---|---|---|
| n | 55 | 80 | 53 | 67 |
| Male, n (%) | 35 (63.6%) | 63 (78.8%) | 34 (64.2%) | 24 (35.8%) |
| Age, Median (Q1, Q3) | 49.5 (40.0, 59.0) | 48.0 (40.0, 59.0) | 60.0 (42.5, 70.0) | 49.0 (34.0, 61.0) |
| Ethnicity, n (%) | | | | |
| - ASIAN | 1 (1.8%) | 1 (1.2%) | 1 (1.9%) | 0 (0.0%) |
| - BLACK/AA | 8 (14.5%) | 12 (15.0%) | 4 (7.5%) | 4 (6.0%) |
| - HISPANIC OR LATINO | 3 (5.5%) | 2 (2.5%) | 1 (1.9%) | 4 (6.0%) |
| - OTHER | 6 (10.9%) | 12 (15.0%) | 7 (13.2%) | 12 (17.9%) |
| - WHITE | 37 (67.3%) | 53 (66.2%) | 40 (75.5%) | 47 (70.1%) |
| Text, n (%) | 29 (52.7%) | 40 (50.0%) | 25 (47.2%) | 27 (40.3%) |
| ICD-9, n (%) | 15 (27.3%) | 40 (50.0%) | 13 (24.5%) | 27 (40.3%) |
| True Positive, n (%) | 36 (66.7%) | 53 (67.1%) | 26 (49.1%) | 37 (55.2%) |

| Variable | Tobacco Use |
|---|---|
| n | 80 |
| Male, n (%) | 46 (57.5%) |
| Age, Median (Q1, Q3) | 54.5 (43.8, 68.2) |
| Ethnicity, n (%) | |
| - ASIAN | 1 (1.2%) |
| - BLACK/AA | 8 (10.0%) |
| - HISPANIC OR LATINO | 2 (2.5%) |
| - OTHER | 7 (8.8%) |
| - WHITE | 62 (77.5%) |
| Text, n (%) | 40 (50.0%) |
| ICD-9, n (%) | 40 (50.0%) |
| True Positive, n (%) | 56 (70.0%) |

### DATA TRANSFORMATION

The data transformation process has been previously described by our group,[1] and is being reproduced here for completeness.

#### Term weighting

A term-by-patient matrix was created where the entries of the matrix were *tf-idf* weighted frequencies of terms across the patient document collection. Weighting (normalization) blunts the effect of common terms while at the same time raising the importance of rare terms that are better discriminators between patients. Each matrix entry $a_{ij}$ is a product of two components: term frequency ($f_{ij}$) and inverse document frequency ($g_i$):

$$g_i = -\log_2 \frac{n_i}{N} \tag{1}$$

where $f_{ij}$ is the frequency of the $i^{\text{th}}$ term in the $j^{\text{th}}$ patient-document, $n$ is the number of patient documents containing the term $i$, and $N$ is the total number of patient documents under consideration.

#### Construction of the LSI model

Singular value decomposition (SVD)[2,3] was applied to the term-by-patient *tf-idf* weighted frequency matrix. Figure S2 demonstrates the process graphically.

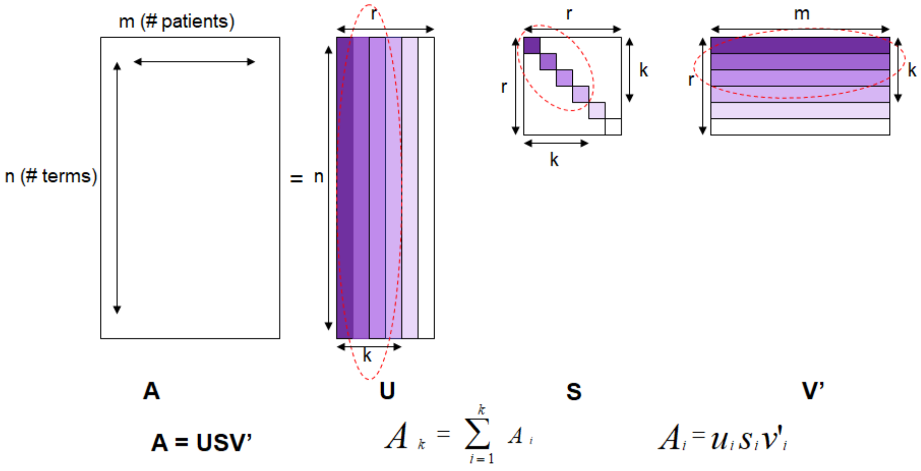


Figure S2: Singular Value Decomposition

A data matrix $A$ with $n$ rows (terms) and $m$ columns (patients), where $n >> m$, can be construed as $n$ term row vectors in $m$-dimensional patient space and $m$ patient-document column vectors in $n$-dimensional term space. SVD transforms the two sets of vectors into a new $r$-dimensional orthogonal space in which the maximum variation is expressed along the first dimension axis, as much variation independent of that is expressed along an axis orthogonal to the first, and so on. The new set of axes may reveal the true dimensionality of the data if the dataset is not inherently $m$-dimensional. The SVD is formulated as:

$$A = USV' \tag{2}$$

where $'$ indicates transpose of the matrix obtained by permuting the modes, i.e., transforming rows into columns and vice versa, $U$ is $n \times r$, $S$ is $r \times r$, and $V$ is $m \times r$ ($V'$ is $r \times m$). Both $U$ and $V$ are orthogonal, i.e., $UU' = I$ and $VV' = I$ where $I$ is the identity matrix. $S$ is a diagonal matrix with non-negative and non-increasing entries $\sigma_1, \sigma_2, \ldots, \sigma_r$ which are known as singular values. $r$ is the rank of the matrix, which is the number of linearly independent rows or columns of $A$. It is however, known from observation, for most practical datasets, $r = m$. The third matrix $V$ is written as a transpose so that the rows of both matrices $U$ and $V$ correspond to terms and patients, respectively.

The rows of $A$ can be interpreted as term coordinates in an $m$-dimensional space. The axes of this space can be interpreted as rows of $I$ (identity matrix). The SVD transforms the term coordinates to rows of $U$ and the axes to the rows of $SV'$. The matrix $V'$ acts as the rotation matrix for the original axes and the diagonal of matrix $S$ contains the scaling factor for each axis. The $U$ matrix can now be construed as a new transformed dataset whose rows still correspond to the original $n$ terms but the patients are transformed into $r$ eigen patients (factors) that are a linear combination of the original patients.

SVD is symmetric in the sense that a decomposition on the rows (terms) can be transformed into a decomposition on the columns (patients):

$$A' = VSU' \tag{3}$$

The new scaled and rotated axes and the coordinates tend to better fit the data than the original axes and coordinates. The singular values in $S$ determine the relative importance of each axis. The first few axes capture the maximum variation in the data and the subsequent ones less so. Only the first $k$ (where $k < r$)

factors corresponding to $k$ largest singular values may be used to represent the data. There are two potential benefits of performing this truncation. Firstly, for large datasets (with many attributes), this translates into savings in memory space as well as analysis time, as vectors in $k$ dimensions can be compared in less time than vectors in $m$ dimensions. Secondly, SVD reveals the true dimensionality present in the data, where the bulk of the information content in the original $m$-dimensional data may be captured in a lower dimensional manifold, after axis rotation and scaling.

##### Calculating most significant factors

An appropriate choice for $k$ (number of most significant factors) can be made by assessing the contribution of each of the singular values as a measure of the amount of variation captured in each dimension, and then calculating the entropy of the contributions that might be indicative of what percentage of the total number of factors may be needed. The contribution $C_i$ of each of $r$ singular values $\sigma$s can be calculated as:

$$C_i = \frac{\sigma_i^2}{\sum_{i=1}^{r} \sigma_i^2} \tag{4}$$

and the entropy of the $r$ contributions calculated as:

$$E = \frac{-1}{\log r} \sum_{k=1}^{r} C_k \log C_k \tag{5}$$

Entropy measures the amount of disorder in the set of variations captured in the $r$ dimensions. The magnitude of the entropy may vary from 0 (all variation is captured in the first dimension) to 1 (all dimensions are equally important). $k$ is calculated as $E \times r$.

The association between any pair of entities (term-term, term-patient, patient-patient) can be calculated as the cosine of the angle between the respective $k$-dimensional vectors. A higher association score between a pair of entities indicates a stronger textual relationship.

#### GPT SBDH Functions

```
housing insecurity
{
        "name": "identify_housing_insecurity",
```

```
        "parameters": {
                "type": "object",
                "properties": {
                        "housing_insecurity": {
                                "type": "string",
                                "enum": ["Yes", "No"]
                        }
                }
        },
        "required": ["housing_insecurity"]
}

financial insecurity
{
        "name": "identify_unemployed",
        "parameters": {
                "type": "object",
                "properties": {
                        "unemployed": {
                                "type": "string",
                                "enum": ["Yes", "No"]
                        }
                }
        },
        "required": ["unemployed"]
}

financial circumstances
{
        "name": "identify_reported_financial_circumstances",
        "parameters": {
```

```
        "type": "object",
        "properties": {
            "reported_financial_circumstances": {
                "type": "string",
                "enum": ["Yes", "No"]
            }
        }
    },
    "required": ["reported_financial_circumstances"]
}

physical or sexual abuse
{
    "name": "identify_history_of_physical_or_sexual_abuse",
    "parameters": {
        "type": "object",
        "properties": {
            "history_of_physical_or_sexual_abuse": {
                "type": "string",
                "enum": ["Yes", "No"]
            }
        }
    },
    "required": ["history_of_physical_or_sexual_abuse"]
}

tobacco use
{
    "name": "identify_tobacco_use",
    "parameters": {
        "type": "object",
```

```
                "properties": {
                        "tobacco_use": {
                                "type": "string",
                                "enum": ["Yes", "No"]
                        }
                }
        },
        "required": ["tobacco_use"]
}

alcohol abuse
{
        "name": "identify_alcohol_abuse",
        "parameters": {
                "type": "object",
                "properties": {
                        "alcohol_abuse": {
                                "type": "string",
                                "enum": ["Yes", "No"]
                        }
                }
        },
        "required": ["alcohol_abuse"]
}

opiate abuse
{
        "name": "identify_opiate_abuse",
        "parameters": {
                "type": "object",
                "properties": {
```

```
                        "opiate_abuse": {
                                "type": "string",
                                "enum": ["Yes", "No"]
                        }
                }
        },
        "required": ["opiate_abuse"]
}

cocaine abuse
{
        "name": "identify_cocaine_abuse",
        "parameters": {
                "type": "object",
                "properties": {
                        "cocaine_abuse": {
                                "type": "string",
                                "enum": ["Yes", "No"]
                        }
                }
        },
        "required": ["cocaine_abuse"]
}

legal circumstances
{
        "name": "identify_reported_legal_circumstances",
        "parameters": {
                "type": "object",
                "properties": {
                        "reported_legal_circumstances": {
```

```
                                "type": "string",
                                "enum": ["Yes", "No"]
                            }
                    }
        },
        "required": ["reported_legal_circumstances"]
}
```
